# Sero-prevalence of visceral leishmaniasis and its associated factors among asymptomatic pastoral community in Denan district, southeastern Ethiopia

**DOI:** 10.1101/2023.01.04.23284183

**Authors:** Ahmed Ismail, Solomon Yared, Sisay Dugassa, Adugna Abera, Abebe Animut, Berhanu Erko, Araya Gebresilassie

**Author notes:** Corresponding author (AG).

## Abstract

**Background:** In the Somali region of Ethiopia, visceral leishmaniasis (VL) is a public health concern. Yet, epidemiology and sand fly vectors of VL were not well studied in various areas of the regional state. Thus, the current study was conducted to determine the sero-prevalence, associated factors and distribution sand fly vectors of VL in Denan district, south-eastern Ethiopia.

**Methods:** A cross-sectional study was conducted among 187 households between May and September 2021 in six selected *Kebeles* of Denan district. In total, 187 blood samples were collected from individuals who visited Denan health center using a simple random sampling technique. Blood samples were subjected to Direct Agglutination Test for the detection of antibodies to VL. Pretested structured questionnaire was used to gather information on individual and household level risk factors and other characteristics of knowledge and attitude assessment. Sand flies were also collected from different sampling habitats using light and sticky traps.

**Results:** The overall sero-prevalence rate was 9.63% (18/187). Sero-prevalence was significantly associated with outdoor sleeping (OR=2.094), presence of damp floor (OR=9.732), and presence of cracked house walls (OR=6.283). Nearly half (53.48%) of the study participants had previously heard about VL. Communities were practicing different methods to prevent VL infection. In total, 823 sand fly specimens comprising 12 species in two genera (*Phlebotomus* and *Sergentomyia*) were trapped and identified. The most abundant species was *S Sergentomyia clydei* (50.18%), followed by *Phlebotomus orientalis* (11.42%).

**Conclusion:** The study demonstrated a relatively higher sero-positivity of VL in this new focus, and a remarkable gap in the knowledge, attitude and practices towards VL. *P. orientalis* was also detected, which could be a probable vector in this new focus. Thus, public education should be prioritized to improve the awareness of the community on VL and its public health impact. In addition, detailed epidemiological and entomological studies are recommended.

## Introduction

Visceral leishmaniasis (VL), also known as Kala-azar, is a vector-borne parasitic disease caused by protozoan parasites of the *Leishmania donovani* complex and transmitted by blood-sucking sand flies. VL is endemic in 75 countries, and the estimated annual global incidence is 50,000–90,000 new cases [1]. About 90% of the global burden for VL is found in just 7 countries, 4 of which are in Eastern Africa (Sudan, South Sudan, Ethiopia and Kenya), 2 in Southeast Asia (India, Bangladesh) and Brazil, which carries nearly all of cases in South America [2,3].

Ethiopia has reported the third largest number of VL cases (1990), following South Sudan (2840) and Sudan (2813) of any country in sub-Saharan Africa region [4]. In Ethiopia, cases of VL have been reported from six regions (Tigray, Amhara, Oromia, Southern Nations and Nationalities People’s Region, Somali and Afar) [5,6], with an annual burden estimated to be between 4, 500 and 7, 400 cases [7]. The incidence rate per 10,000 people in endemic areas is 6.28 [4]. This systemic disease is known to be endemic in Metema and Humera plains in northwest; in several localities of south western Ethiopia, i.e., the Omo plains, the Aba Roba focus in Segen valley, and Woito River valley adjacent to South Omo; in southern Ethiopia around Moyale area close to the borders with North Kenya; and in south eastern Ethiopia around Genale river basin in Oromia Regional State, Afder and Liban zones in Somali Regional State [6].

In the Somali Regional State, VL outbreaks were first reported in 2001 from Afder, Liben, Denan and Hagele areas, bordering Kenya and Somalia [8]. Subsequently, VL cases have been reported sporadically from different areas of the regional state [9,10]. What is more, the national risk map survey indicated that a wide geographical areas stretching from south-eastern part of the Somali region to the Ethio-Somali-Kenyan boundaries as high-risk areas for VL [11]. Despite the public health significance and the recent occurrences of VL in different foci of the regional state, epidemiology and risk factors of VL were not adequately addressed.

Following continuous collections of sand flies to determine their fauna and epidemiological significance in the transmission of leishmaniases in different parts of the Ethiopia, so far 65species belonging to the genus *Phlebotomus* and *Sergentomyia* have been identified [12]. Of these, the incriminated vectors of VL, from which parasites have been detected and/or isolated and identified, include *P. martini, P. celiae*, and *P. orientalis* for *L. donovani* from the south, south-west and northern foci [13,14,15]. Study on sand fly vectors in VL endemic foci is very important to improve the understanding of the transmission dynamics and design vector control methods to reduce the burden of the disease. However, there are gaps in the knowledge of habitat preferences, behavior and distribution of sand flies in various VL foci of the Somali region.

Therefore, the investigation was designed to determine the sero-prevalence, knowledge, attitude and practices, and associated factors of VL, and abundance of sand fly species in Denan district, south-eastern Ethiopia.

## Materials and Methods

### Study Area

The study was carried out in Denan district of Shebelle zone in south-eastern Ethiopia (Fig 1). Denan town, the administrative center of the district, is located 1,123 kilometres southeast of Addis Ababa. It is found at an elevation of about 490 meters above sea level, with a latitude and longitude of 6° 44’ 59.99” N and 43° 19’ 60.00” E, respectively. Denan has a subtropical desert climate with yearly temperature of 29.39ºC. It typically receives about 19.24 millimeters of precipitation annually. The total population of Denan district is about 33,784 people according to the last Ethiopian national census of 2007 [16]. The district is composed of 12 administrative *kebeles* (the smallest administrative unit). Of which, six kebeles (Qora, Kore, Shinile, Birta-dheer, Awley, and Burqeyr) were selected for this study. Denan district has two health centres and nine health posts.

**Figure 1:**
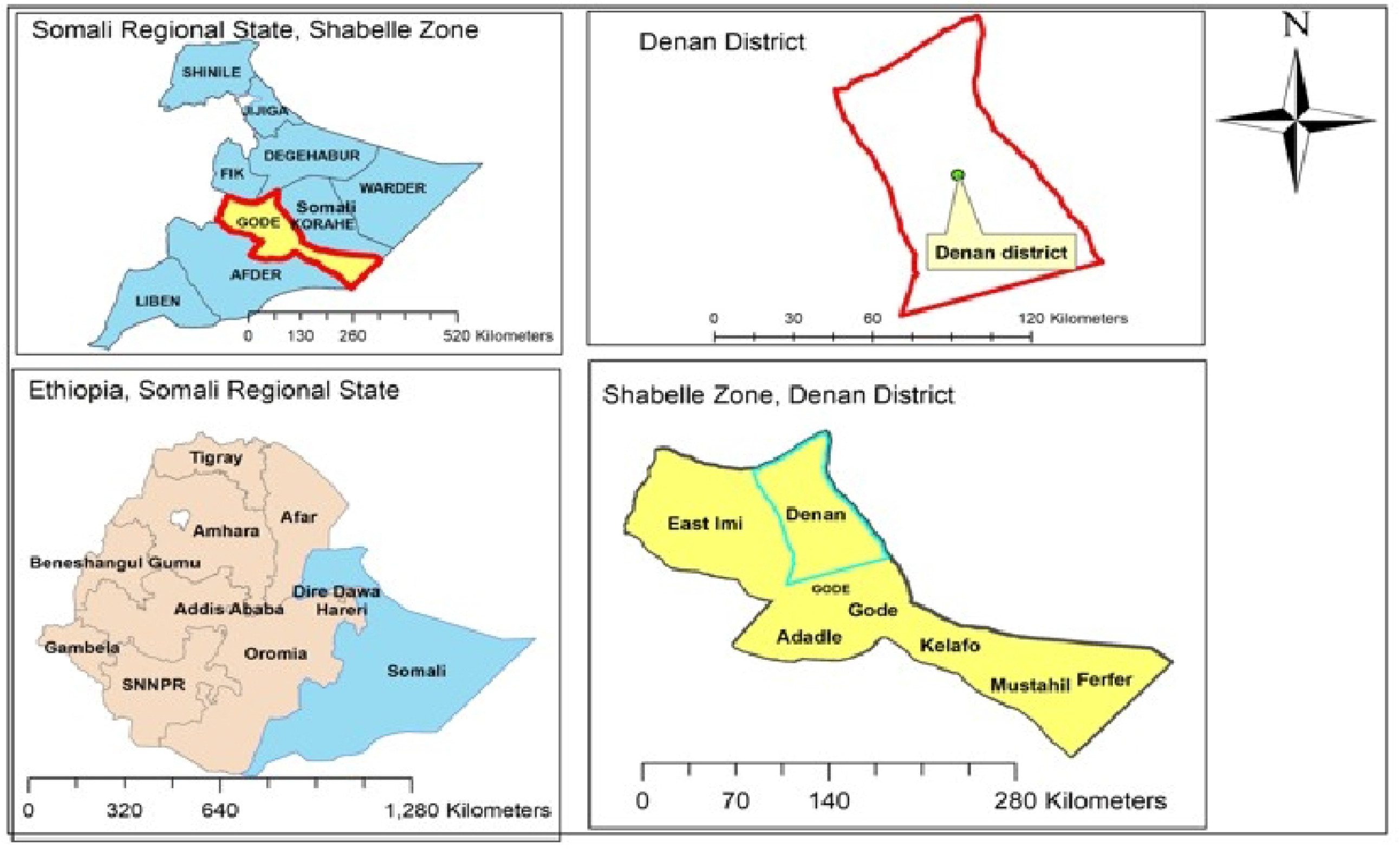
Map of the study area in Shebelle zone of Somali Regional State, southeastern Ethiopia.

### Study design and sample size determination

A cross sectional study design was employed to determine the sero-prevalence of VL and identify the associated risk factors from April to August 2021. The sample size was estimated according to [17] by considering 15.8% previous prevalence of VL in a nearby district [10], 95% confidence interval (CI) (Z=1.96) with 5% desired absolute precision (d), using the formula N=[(Z)^2^ X P (1-P)]/d^2^. Consequently, a total of 204 human sera were required, however, 17 participants were excluded from the study as they refused to give blood sample.

### Sampling procedure

Permanent residents and pastoral communities of the selected sites were enrolled in the study. Individuals who previously had VL were excluded. Single stage random sampling was applied for selecting the study participants for the field survey. Six *kebeles* including Qora, Kore, Shinile, Birta-dheer, Awley, and Burqeyr were selected. The sampling *kebeles* were selected based on the suspected cases of VL by reviewing case records of Denan health center, and transport accessibility. Households (187) were randomly selected from the six *kebeles*. One participant was then randomly selected from each household and contacted for informed consent/assent. The participants were requested to come in to Denan health center and in a nearby health posts for blood sample collection.

### Blood Sample Collection

Five ml of venous blood was collected following standard operating procedures. Blood samples were obtained by skilled laboratory technician from the forearm veins of the study participants using sterile needle. The blood sample was dispensed into serum separator test tubes at Denan Health Centre lab and transported in a cool box to Addis Ababa. The serum was diagnosed by DAT at Ethiopian Public Health Institute.

### Direct Agglutination Test (DAT)

In performing DAT, serum samples were serially diluted in physiological saline (0.9% NaCl) containing 0.8% β-mercaptoethanol. Two-fold serial dilutions of the sera were made, starting at a dilution of 1:100 and going up to a maximum serum dilution of 1:102,400. Freeze-dried DAT antigen produced by KIT Biomedical Research (DAT, Institute of Tropical Medicine—Antwerp (ITM), Belgium) was reconstituted with physiological saline. 50 μL of DAT antigen solution (concentration of 5 × 10^7^parasites per ml) was added to each well containing 50 μl of diluted serum. The results were read after 18 hours of incubation at ambient temperature. The cut-off value was established considering the titres obtained in samples from negative controls. Therefore, a sample was considered positive if it had a titre of 1:800 and above.

### Assessment of KAP and associated factors

A pre-tested structured questionnaire was designed to collect information regarding socio-demographics and associated factors such as house construction material and its condition, domestic animal ownership, sleeping habits, use of bed net and individual night and day activity. The questionnaire also comprised of questions relating to the respondents’ knowledge, attitudes and common practices towards VL and sand flies. The questionnaire was first developed in English and translated into Af-Somali (the local language). Data were checked for completeness, and incomplete questionnaires were returned to data collectors for correction by revisiting the concerned households.

### Sand fly collection and processing

Entomological investigations were undertaken in four *kebeles* (Kore, Shinile, Birta-dheer, Awley) of Denan district during 4-12 April, 2021. The above four kebeles were selected as sampling sites based on suitability of habitats for species richness and abundance of sand flies and accessibility to transport. In the sampling villages, four representative trapping habitats such as indoor, peri-domestic, mixed forest and termite mounds were identified and used for the entire sand fly species collection. Sand flies were trapped for two consecutive nights at each sampling village, constituting a total of 12 collection nights. Sand flies were collected using CDC light traps and sticky traps.

#### CDC light traps (CDC-LTs)

LTs were deployed at peri-domestic, mixed forest and termite mounds. Two LTs were set in representative sites of peri-domestic habitats. Another two LTs in each village were positioned to sample sand flies in mixed forest. Similarly, LTs were suspended in termite mounds. The traps were deployed 1 h before sunset and collected at dawn the next morning. Afterwards, the sand flies were sorted by sex and genus (*Phlebotomus* or *Sergentomyia* spp.), and preserved in 70% ethanol for later species identification.

#### Sticky traps (ST)

A4-sized white sticky traps of polypropylene sheets coated with sesame oil were used for capturing sand flies from all sampling habitats. The ten STs were divided into 2 sets each having five sheets tied together on nylon string about 50 cm apart and these were placed inside two different houses in each village to intercept and capture any endophilic sand flies. Similarly, two sets of sticky traps were also suspended randomly on cracked walls, stone piles, and animal enclosures in the peri-domestic environment. Another four sets of STs were also deployed in representative sites of mixed forest and termite mounds. Each morning, sand-flies from STs were removed using forceps and stored in 96% ethyl alcohol in labeled vials for identification.

### Mounting and identification of sand flies

Sand fly specimens were dissected and mounted on microscope slides in Hoyer’s medium with their heads separate from thoraxes and abdomens. Slide-mounted flies were then identified to species level based on the external genitalia of males and the pharynx, antennal features and spermathecae of females, according to standard morphological keys [18].

### Data analysis

Statistical analyses were conducted using IBM SPSS statistics, version 20 for Windows (SPSS Inc., Chicago, IL, USA), and Microsoft® Office Excel 2007. Descriptive statistics was computed to determine frequency and percentage. The Chi square statistic was used to determine the associations between socio-demographic characteristics and VL positivity. Logistic regression was used to determine possible factors associated with VL. For all included studies, a P-value < 0.05 was regarded as statistically significant.

### Ethical consideration

This study was conducted after the protocol was reviewed and approved by the Institutional Review Board (IRB) of Aklilu Lemma Institute of Pathobiology, Addis Ababa University. Permission to conduct the study was also obtained from the Denan district health office after having thorough discussion on the procedures and purpose of the study. In addition, verbal informed consent was obtained from all selected individuals for the study after explaining the purpose of the study in the local language, Af-Somali.

## Results

### Socio-demographic characteristics

A total of 187 participants recruited from six *kebeles* of Denan district were included. Out of the total respondents, 119 (63.64%) and 68 (36.36%) were males and females, respectively. The minimum age of an individual included in the study was 18 years. The majority of respondents [110 (58.82%)] had no formal education. Most of the respondents [110 (54%)] had family size of 3. Majority of the participants were pastoralists (54.55%). Higher proportions of participants belonged to households living in Aqal, dome-shaped huts of traditional Somali house, (55.1%), with abundant termite mounds (48.1%) (Table 1).

**Table 1:** Socio demographic characteristics of study participants of Denan district, Somali Region, south east Ethiopia, 2021.

| Variable | Categories | Frequency | Percentage |
| --- | --- | --- | --- |
| Age | >18 | 44 | 23.53 |
|  | 19-36 | 108 | 57.75 |
|  | 37-54 | 27 | 14.44 |
|  | ≥55 | 8 | 4.28 |
| Sex | Male | 119 | 63.64 |
|  | Female | 68 | 36.36 |
| Marital status | Single | 58 | 31.02 |
|  | Married | 127 | 67.91 |
|  | Widowed | 2 | 1.07 |
| Family size | ≤ 3 | 101 | 54 |
|  | ≥ 4 | 86 | 46 |
| Community lifestyle | Pastoralist | 102 | 54.55 |
|  | Agro-pastoralist | 23 | 12.3 |
|  | Urban | 62 | 33.16 |
| Education | Illiterate | 60 | 32.09 |
|  | Religious education | 50 | 26.74 |
|  | Primary | 37 | 19.79 |
|  | Secondary schooling and above | 40 | 21.39 |
| House type (wall) | Woven mats (Aqal) | 103 | 55.10% |
|  | Mud and stick | 43 | 22.90% |
|  | Concrete blocks | 41 | 21.90% |
| Termite mounds near house | Yes | 90 | 48.10% |
|  | No | 97 | 51.90% |
| Total |  | 187 |  |

### Sero-prevalence and associated factors of VL

The overall sero-prevalence rate of VL in the study area was 9.63% (18/187). Sero-prevalence rates did not significantly vary between males (6.4%) and females (3.2%) (P>0.05, Table 2). Individuals in the age groups 19-36 and 37-54 had highest DAT positivity rates of 5.9% and 2.1%, respectively, while the lowest positivity rates were observed within the age group of less than 55 (0.5%), followed by less than equal to 18 (1.1%) (Table 2). The difference in DAT positivity by age group was statistically significant (P<0.05).

**Table 2:**
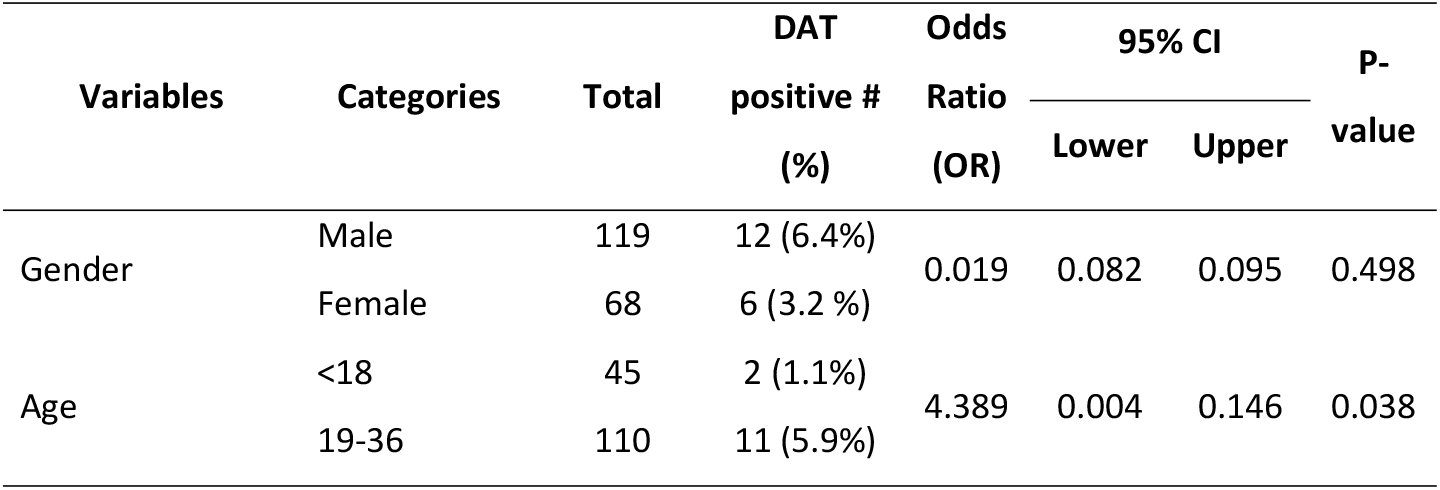
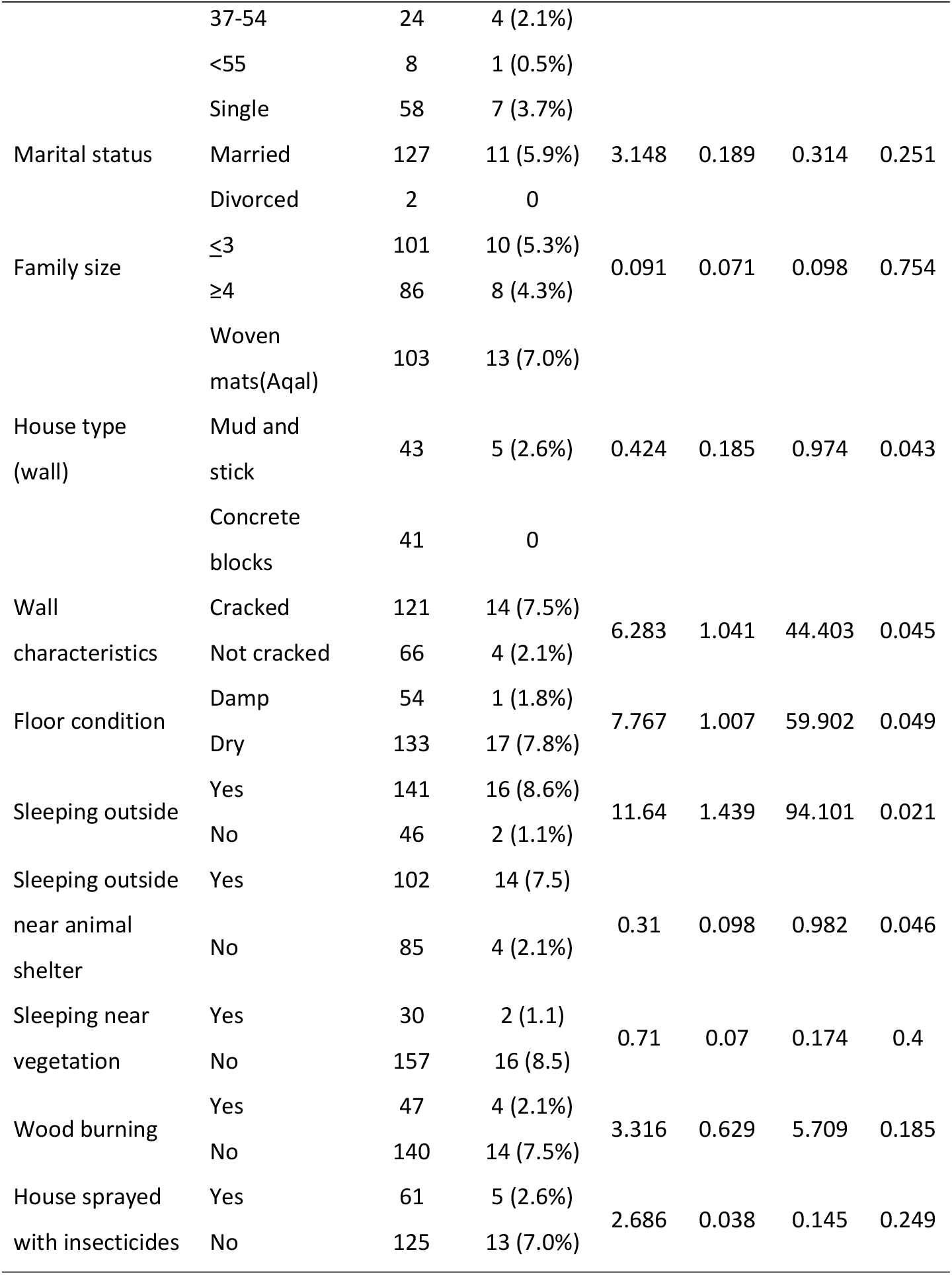
Factors associated with transmission of visceral leishmaniasis in univariate analysis in Denan district, Somali Region, southeast Ethiopia, 2021.

Significant differences in the prevalence of VL were found among age groups, housing type (walls), floor condition, outdoor sleeping, and sleeping near animal shelter (P<0.05). However, variables such as sex, family size, marital status, wood burning, sleeping near specific vegetation, and house spray with insecticide were found to be statistically non-significant (P>0.05) (Table 2).

Table 3 shows risk factors associated with VL in multivariable logistic regression models. Being in the age group of 19-36 increases the odds of getting VL infection (P=0.034). Similarly, individuals with the habit of sleeping outdoors near animal shelter likely to be at risk of acquiring VL infection than those sleeping indoor. Participants who owned houses with damp floor were found to have high DAT positivity compared to those who had their house with dry floor (Table 3).

**Table 3:** Factors associated with transmission of VL in multivariate analysis in Denan district, Somali Region, southeast Ethiopia, 2021.

| Variables | Categories | Total | DAT<br>positive #<br>(%) | Odds<br>Ratio<br>(OR) | 95% CI |  | P-value |
| --- | --- | --- | --- | --- | --- | --- | --- |
|  |  |  |  |  | Lower | Upper |  |
| Age | >18 | 45 | 2 (1.1%) | 0.441 | 0.207 | 0.94 | 0.034 |
|  | 19-36 | 110 | 11 (5.9%) |  |  |  |  |
|  | 37-54 | 24 | 4 (2.1%) |  |  |  |  |
|  | ≤55 (ref) | 8 | 1 (0.5%) |  |  |  |  |
| Floor condition | Dry (ref) | 54 | 1 (1.8%) | 0.09 | 0.012 | 0.705 | 0.022 |
|  | Damp | 133 | 17 (7.8%) |  |  |  |  |
| Outdoor sleeping | Yes | 141 | 16 (8.6%) | 4.35 | 2.052 | 9.224 | 0.001 |
|  | No (ref) | 46 | 2 (1.1%) |  |  |  |  |
| Sleeping outdoor<br>near animal shelter | Yes | 102 | 14 (7.5) | 2.521 | 0.534 | 11.908 | 0.031 |
|  | No (ref) | 85 | 4 (2.1%) |  |  |  |  |

### Knowledge of VL and sand fly vectors

Among the total participants, 100 (53.48%) had heard about the disease, and 42% responded that sand fly bite is responsible for VL transmission (Table 4). The most common sources of VL related information were health personnel (48%), followed by friends and neighbors (39%), mass media (radio and television) (5%), and school (8%). Most of the respondents (89%) knew at least one clinical sign and symptom of VL, and 41% of them indicated that abdominal swelling is the key symptom of VL. A substantial number of respondents (78%) mentioned the disease as treatable. Around 87.5% of respondents replied that they were aware of sand fly breeding grounds, and 21% indicated that they have multiple breeding habitats.

**Table 4:** Knowledge on VL among study participants Denan district, Somali Region, southeast Ethiopia, 2021.

| Variables | Categories | Frequency | Percentage (%) |
| --- | --- | --- | --- |
| Heard about VL (n = 187) | Yes | 100 | 53.48 |
|  | No | 87 | 46.52 |
| Source of VL information (n=100) | Health personnel | 48 | 48 |
|  | Friends and neighbors | 39 | 39 |
|  | Media (Radio and TV) | 5 | 5 |
|  | School | 8 | 8 |
| Mode of transmission (n=100) | Contact with sick people | 18 | 18 |
|  | Sandfly bite | 42 | 42 |
|  | Spoiled food | 3 | 3 |
|  | Mosquito bite | 23 | 23 |
|  | Don't know | 14 | 14 |
| VL signs and symptoms | Fever | 7 | 7 |
|  | Headache and fatigue | 3 | 3 |
|  | Abdominal swelling | 41 | 41 |
|  | Multiple answers | 38 | 38 |
|  | Don't know | 11 | 11 |
| Treatability of VL (n=100) | Yes | 78 | 78 |
|  | No | 7 | 7 |
|  | Don't know | 15 | 15 |
| Identify the sand fly (n=187) | Yes | 96 | 51.33 |
|  | No | 91 | 48.7 |
| Sand flies found around your houses | Yes | 76 | 79.17 |
|  | No | 20 | 20.83 |
| Sand fly breeding sites | Cracks of houses | 20 | 20.8 |
|  | Termite mounds | 14 | 14.6 |
|  | Near animal shelter | 12 | 12.5 |
|  | Crevices of tree | 17 | 17.7 |
|  | Multiple domestic areas | 21 | 21.9 |
|  | Don't know | 12 | 12.5 |

### Attitude and practices of the participants on VL and sand flies vector

Table 5 shows attitudes and practices of the participants about visceral leishmaniasis. Below 50% of the respondents stated that VL is an important public health problem in the area, while 52% considered the opposite. Majority (68%) of the respondents preferred public health care services as their first priority for VL treatment. Regarding the fatality, 44% of the respondents claimed that VL is fatal at all. Only 36% of the respondents believed that VL is preventable (Table 5).

**Table 5:**
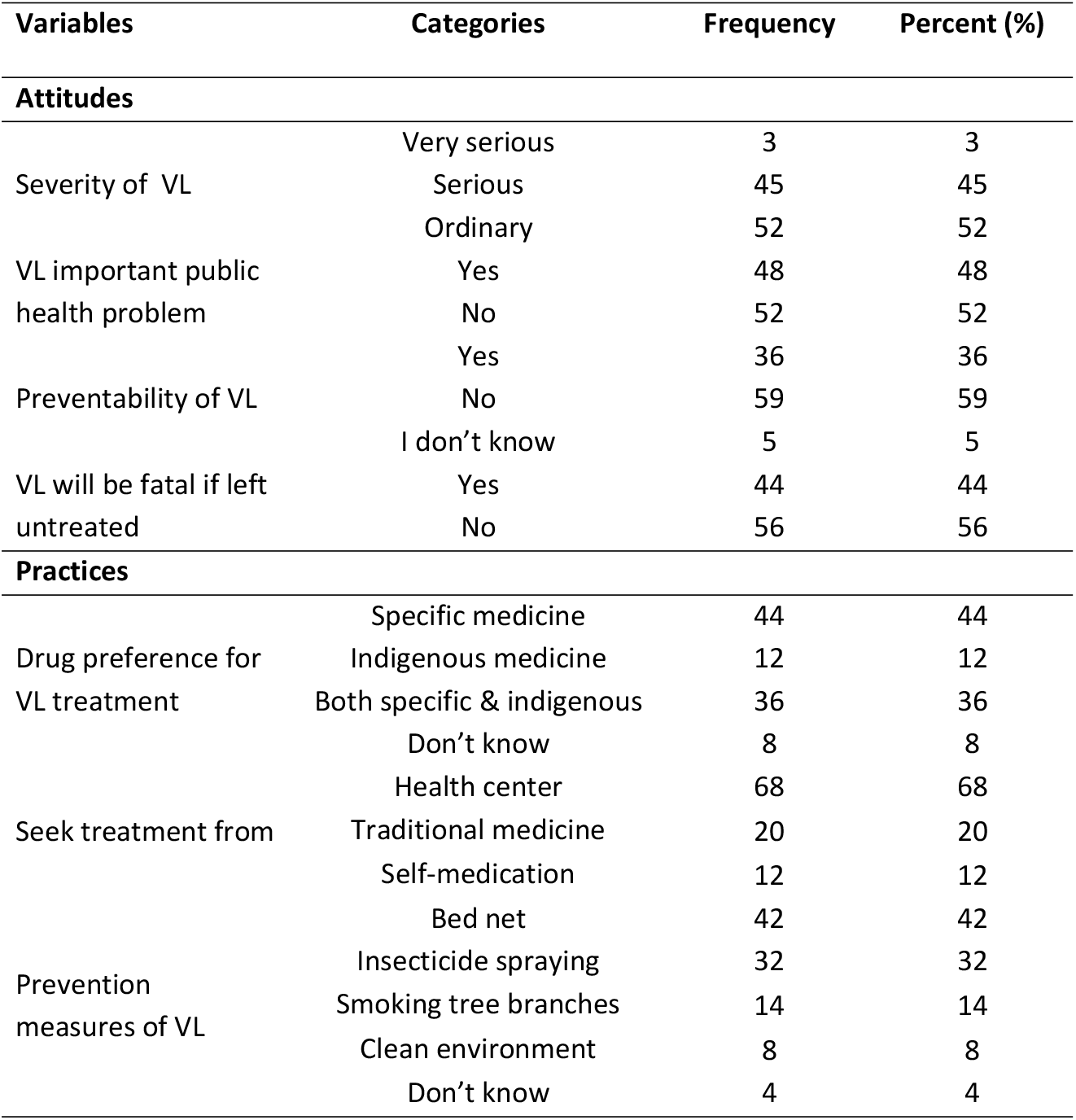
Attitude and practice of the participants on VL and sand flies vectors (n= 100)

In this study, participants were practicing different methods to prevent VL: 42% of the respondents used a bed net, 32% used insecticide spraying, 14% practiced smoking plant parts, and 8% cleaned their environments (Table 5).

### Species composition of trapped sand fly

A total of 823 sand flies were collected and identified. The sand fly fauna included both the genus *Phlebotomus* and *Sergentomyia* (Table 6). Twelve species were morphologically identified to the species level, and these sand fly species were *P. orientalis, P. alexandri, P. papatasi, P. rodhaini, P. saevus, S. clydie, S. bedfordi* group, *S. schwetzi, S. africana, S. antennata, S. squamipluris*, and *S. heisch* (Table 6). In total, 355 (43.13%), 284 (34.51%), 174 (21%), and 10 (1.22%) sand fly specimens captured using LTs and STs deployed in termite mounds, forest, peri-domestic, and indoor habitats, respectively (Table 6).

**Table 6:** Species of sand flies collected from different sampling sites of Denan district, Somali Region, southeast Ethiopia, 2021.

| Species | Number of sand fly specimens in different sampling habitats |  |  |  |  |  |  |  | Total |
| --- | --- | --- | --- | --- | --- | --- | --- | --- | --- |
|  | Peri-domestic |  | Indoor |  | Forest |  | Termite mounds |  |  |
|  | Male | Female | Male | Female | Male | Female | Male | Female |  |
| *Phlebotomus* |  |  |  |  |  |  |  |  |  |
| *orientalis* | 5 | 5 | 0 | 0 | 13 | 18 | 29 | 24 | 94 |
| *P. alexandri* | 2 | 6 | 1 | 1 | 19 | 21 | 6 | 9 | 65 |
| *P. papatasi* | 9 | 19 | 0 | 1 | 4 | 3 | 3 | 7 | 46 |
| *P. rodhaini* | 0 | 1 | 0 | 0 | 4 | 0 | 0 | 0 | 5 |
| *P. saevus* | 0 | 1 | 0 | 0 | 0 | 0 | 2 | 1 | 4 |
| *Sergentomyia* |  |  |  |  |  |  |  |  |  |
| *clydie* | 41 | 40 | 6 | 0 | 72 | 59 | 111 | 84 | 413 |
| *S. bedfordi* group | 1 | 13 | 0 | 0 | 19 | 26 | 11 | 20 | 90 |
| *S. schwetzi* | 1 | 3 | 0 | 1 | 2 | 3 | 18 | 16 | 44 |
| *S. africana* | 0 | 14 | 0 | 0 | 3 | 13 | 4 | 6 | 40 |
| *S. antennata* | 2 | 2 | 0 | 0 | 2 | 3 | 3 | 0 | 12 |
| *S. squamipluris* | 5 | 4 | 0 | 0 | 0 | 0 | 0 | 0 | 9 |
| *S. heischi* | 0 | 0 | 0 | 0 | 0 | 0 | 1 | 0 | 1 |
| **Total** | 66 | 108 | 7 | 3 | 138 | 146 | 188 | 167 | 823 |

### Relative abundance and sex ratio of sand fly species

Of the 214 *Phlebotomus* specimens, *P. orientalis* constituted 43.9% of sand flies captured, followed by *P. alexandri* (30.37%), *P. papatasi* (21.49%), *P. rodhaini* (2.33%), and *P. saevus* (1.86%). Among *Sergentomyia* spp., *S. clydei* was the most predominant species, accounting for 67.81% and 50.18% of *Sergentomyia* species and the entire sand fly collection, respectively. Abundance of other *Sergentomyia* species in descending order were *S. bedfordi* group (14.78%), *S. schwetzi* (7.22%), *S. africana* (6.57%), *S. antennata* (1.97%), *S. squamipluris* (1.48%), and *S. heischi* (0.16%) (Table 7).

**Table 6:** Relative abundance and sex ratio of sand fly species from Denan district, Somali Region, southeast Ethiopia

| Species | Type of collections methods |  |  |  |  | Sex ratio | Total | Relative frequency (%) |
| --- | --- | --- | --- | --- | --- | --- | --- | --- |
|  | Light traps |  | Sex ratio | Sticky traps |  |  |  |  |
|  | M | F |  | M | F |  |  |  |
| *P. orientalis* | 25 | 28 | 0.89 | 22 | 19 | 1.16 | 94 | 11.42 |
| *P. alexandri* | 17 | 25 | 0.68 | 11 | 12 | 0.92 | 65 | 7.89 |
| *P. papatasi* | 11 | 24 | 0.46 | 5 | 6 | 0.83 | 46 | 5.59 |
| *P. rodhaini* | 1 | 1 | 1 | 3 | 0 | NA | 6 | 0.61 |
| *P. saevus* | 1 | 2 | 0.5 | 1 | 0 | NA | 4.5 | 0.47 |
| *S. clydie* | 102 | 113 | 0.90 | 128 | 70 | 1.83 | 413 | 50.18 |
| *S. bedfordi group* | 14 | 36 | 0.39 | 17 | 23 | 0.74 | 90 | 10.94 |
| *S. schwetzi* | 5 | 4 | 1.25 | 16 | 19 | 0.84 | 45 | 5.35 |
| *S. africana* | 4 | 28 | 0.14 | 3 | 5 | 0.6 | 40 | 4.865 |
| *S. antennata* | 4 | 5 | 0.8 | 3 | 0 | NA | 12 | 1.46 |
| *S. squamipluris* | 5 | 4 | 1.25 | 0 | 0 | NA | 10 | 1.096 |
| *S. heischi* | 0 | 0 | 0 | 1 | 0 | NA | 1 | 0.126 |
| **Total** | 189 | 270 | 0.7 | 210 | 154 | 1.36 | 823 |  |

Overall male/female ratio for different species of sand flies was 0.94 (Table 7), with 51.5% females and 48.5% males. In light traps, the sex ratio was 0.7, while on the sticky traps it was slightly in favor of males with 1.36:1.

## Discussion

Wide areas in the Somali region have been predicted to have a high VL risk based on the environmental factor-based geographical information and statistics risk mapping [11]. Nevertheless, data on the epidemiology of VL in various endemic areas of the region barely exist. In this regard, a study was carried out to determine the sero-prevalence, associated factors and sand fly vectors of VL in Denana district, south-eastern Ethiopia. In this study, the overall sero-prevalence of *L. donovani* among asymptomatic individuals was 9.63% by DAT. The DAT positivity rate of 9.63% in this survey was lower than previous rate of 12.5% among migrant laborers in Kafta-Humera lowlands [19], and higher than that from Raya Azebo, northeastern Ethiopia [20], and from different areas of Benishangul-Gumuz region, western Ethiopia [21], who reported 0.8% and 5.9%, respectively. In addition, the current sero-prevalence rate was higher than previous report of 3.0% using ELISA test from neighboring districts of Gode and Adadle, southeastern Ethiopia [10]. Possible reasons for such variation could be related to difference in agro-ecological settings, possible predisposing risk factors, serological diagnostic techniques, and intervention strategies.

Understanding factors that determine VL exposure in endemic foci provide a vital foundation of knowledge for designing and developing effective control methods. The findings of this study showed that adults in the age group 19-36 had elevated VL risk. In contrast to this finding, earlier studies in Ethiopia [10,22,23], and elsewhere [24,25,26] have shown a higher risk of VL in individuals under 15 years compared to adults. The contributing factors for such higher burden of disease among adults might be related to activities like migratory and outdoor lifestyles, involving cattle herding or sleeping outside, that imply an increased potential exposure to the sand fly vector, and that are culturally specific to male adolescents and male adults. The differences in design may also account for this difference, as the cited studies were carried out in a population with a different age distribution with more than 50% of the sample being below 15 years.

The results also showed that there is a strong association between VL and poor housing condition such as damp floor. Floor dampness was identified as a major independent risk factor in India and Nepal [27,28], apparently showing that houses with damp floors could provide the adequate needs for the sand fly vector survival. Outdoor sleeping was identified as a risk factor for VL in our study area, which is also consistent with other reports [10,23,29,30]. However, owning large family size was not found to be associated with increased VL risk in our study area, which is in disagreement with earlier reports that large family size was associated with a higher risk of VL [31,32].

The current study also assessed participants’ knowledge, attitude, and practices about VL infection. A little more than half of the respondents (53.48%) had heard about VL. Our finding is lower than the 85% and above found in earlier reports in Ethiopia [33]. Only 42% of study participants knew that sand fly is the cause of VL transmission. This value is higher than the findings of reports on VL from Bihar in India (32.1%) [34], and northwest Ethiopia (30%) [32]. However, this result is lower than the 68.1% found in a study in northwest Ethiopia [33]. Moreover, most of the respondents (89%) knew at least one clinical sign and symptom of VL. This figure is in agreement with a report from Welkait district, northwest Ethiopia, where 88% of the respondents were aware of at least one clinical sign [35]. Nevertheless, our value exceeds the findings of a report on VL in northwest Ethiopia (62%) [33]. Such variability of knowledge among the various studies could be associated with the difference in the setting of the study and public awareness activities conducted, and the fact that VL and its related information is not well established in the area.

Most of the respondents (59%) believed that VL is not preventable, while only 36% of the study participants said that the disease could be prevented. Our result is in contrast with Alemu et al. [33] and Berhe et al. [35], who reported that 81.2% and 90.5% of respondents believe VL is treatable. In addition, only 44% of the study participants knew that if the disease is left untreated the outcome will be death, which is by far lower than that of a study reported from northwest Ethiopia [33], where 96.7% of participants perceive that VL is fatal. In our study area people have poor perceptions towards VL preventability and its fatality, suggesting more has to be done to increase the awareness of the community.

In this study, it was evidenced that on top of bed net, other preventive and control activities were practiced by the study participants to protect themselves from any biting flies including sand flies. Forty two percent of the respondents use bed nets, 32% insecticide spraying, 14% smoking tree branches, and 8% clean their environments against for the control of VL transmission by sandfly bites. This percentage is higher than that of a study carried out in the Indian state of Bihar [34]; where 21.4% of the respondents used bed nets for the control of sand fly bites. Nevertheless, it is lower than a report from northwest Ethiopia, where around 70% of the respondents used a bed net for the prevention of VL [32]. Awareness creation and sensitization activities performed in relation to malaria control, the availability of anti-mosquito repellants in most drug shops and culture of the community to use plant parts for different smoking activities may have contributed to better practices towards the control and prevention of VL in our study area.

In general, this study indicated that the overall unfavorable knowledge and attitude about the disease was alarming in a sense far from satisfactory. In addition to that, specific habitats of sandfly resting and breeding areas were seen in all the study kebeles. On top of that, residents in the study area practiced outdoor sleeping activities owing a very high risk for being a blood meal source for exophilic vectors species in these ecological settings.

Along the sero-prevalence and KAP studies, an entomological investigation was carried out to determine the sand fly vectors of VL. Among the 65 sand fly species known in Ethiopia, 12 (18.5%) species of sand flies were identified in this study. The sand fly species composition recorded in the this study is concurrent with previous reports in other parts of Ethiopia [10,36-38]. Five species of *Phlebotomus* were identified during this study, where *P. orientalis* was the dominant species, constituting 11.42% of total sand fly captures. *P. orientalis* is the vector of VL caused by *Leishmania donovani* in Sudan, South Sudan [39,40], southwestern Ethiopia [13], and the northern Ethiopia [15]. Apparently, this sand fly species could be involved in the transmission of VL in this new VL focus. As a result, further studies to determine the infection rates and host preference patterns of this vector species are required.

Another epidemiologically important species of the genus *Phlebotomus* found during our surveys included *P. alexandri, P. papatasi, P. rodhaini, and P. saevus*. The vectorial status of *P. alexandri* is not clearly established in different parts of Ethiopia despite it is a proven vector of *L. infantum*, causative of zoonotic VL, in China [41]. *P. rodhaini* reported in this study is also suspected as a VL vector in eastern Sudan and is implicated in maintaining the zoonotic cycle between reservoir animals [42]. *L. tropica* was also isolated from two specimens of *P. saevus* in the Awash Valley, northeastern Ethiopia [43]. Similarly, *P. papatasi* is the principal vector of *L. major* in most parts of the Old World [44], albeit its vectorial role in CL transmission in Ethiopia is yet unclear.

Species of the genus *Sergentomyia* were almost predominant in the collections compared to the genus *Phlebotomus* with 74.24% of species identified. Such preponderance has already been observed in many surveys, notably in Ethiopia and elsewhere in Afrotropical regions [45,46].

Regarding the microhabitat preference, most of the *P. orientalis* were captured from termite mounds followed by forest dwellings. Such higher abundance of this species in outdoor habitats could be related to the greater availability of suitable resting and breeding habitats. In addition, some of the vegetation could provide shade and source of sugar for adult populations [47]. However, no single specimen of *P. orientalis* was caught indoor, which is in agreement with earlier study in southeastern part of the Somali region [38].

## Conclusions

In conclusion, the sero-prevalence of VL among the asymptomatic pastoralist community is relatively higher as new VL focus. Adults in the age group 19-36 showed higher VL sero-reactivity compared to other age groups in the study. The main factors associated with increased risk of VL infection factors were living in houses with damp floor, outdoor sleeping behaviors, and sleeping near animal shelter. Poor knowledge coupled with unfavorable attitudes and practices towards the disease are also alarming issues against the ease to control the disease, suggesting the need to increase awareness of VL transmission and prevention in pastoral communities. *P. orientalis*, proven vector of VL, apparently contributes to the VL transmission in this new VL focus. Further wide-ranging studies on the epidemiology and magnitude of the disease and investigations of other contributing risk factors in the area are required. Concurrently, additional studies to understand the bionomics of *P. orientalis* should be carried out to effectively control the disease.

## Data Availability

All data produced in the present study are available upon reasonable request to the authors

## Acknowledgements

The authors would like to acknowledge Addis Ababa University for sponsoring the study. We also thank Ethiopian Public Health Institute for allowing us to use the lab facility to run DAT. Our appreciation also goes to Denan District Health Office for or their valuable information and support.

## Author contributions

Conceptualization: Ahmed Ismail, Solomon Yared, Araya Gebresilassie

Data curation: Ahmed Ismail, Araya Gebresilassie

Formal analysis: Ahmed Ismail, Solomon Yared, Sisay Dugassa, Abebe Animut, Berhanu Erko, Adugna Abera, Araya Gebresilassie

Methodology: Ahmed Ismail, Solomon Yared, Adugna Abera, Araya Gebresilassie

Writing – original draft: Ahmed Ismail, Solomon Yared, Araya Gebresilassie

Writing – review & editing: Ahmed Ismail, Solomon Yared, Sisay Dugassa, Abebe Animut, Berhanu Erko, Adugna Abera, Araya Gebresilassie

